# Identification of a Fractional Model for an Outbreak of the Dengue Fever

**DOI:** 10.64898/2026.05.26.26354120

**Authors:** Jacky Cresson, Marielle Péré, Anna Szafrańska

## Abstract

This work focuses on the global and partial identification problem for fractional differential equations. We provide a general numerical procedure based on global and local optimization algorithms with two refinements for biological systems that ensure solution positivity and homogeneous parameter units. The method is applied to a new fractional model of Dengue outbreak called the Fractional Homogeneous Nishiura (FHN) model, calibrated using data of newly infected people in Cape Verde.

We show that our identification method yields a better fit between data and model solutions than previous approaches and that our FHN model captures the dynamics of Dengue more closely than existing systems.

## 1 Introduction

Partial observability is the norm in complex dynamical systems, particularly in life sciences. While mechanistic models are becoming increasingly complex to capture both intra-process dynamics, impact of the environment and potential adaptation to stress and perturbations, calibration data on natural phenomena remain noisy, limited and often delayed, reflecting only a fraction of the true system state. Process monitoring but also the time evolution of the process itself modify the data quality over time. This mismatch between model complexity and data quality creates a fundamental parameter identification problem that prevents the implementation of early predictive control measures in many cases, such as in epidemiology. To address the challenge of model complexity, fractional differential equations (FDEs) are a powerful mathematical framework to refine and extend ordinary differential equation models of natural phenomena [11] without multiplying the number of free parameters.

Fractional differential equations (FDEs) extend the classical notion of differentiation to non-integer orders, providing a natural framework for modeling complex systems exhibiting memory effects and non-local interactions [29]. These properties make FDEs particularly well-suited for capturing the dynamics of social and biological phenomena like disease outbreaks, whose non-Markovian nature and dependence on past states are poorly represented by classical integer-order models. However, despite introducing a relatively moderate number of new parameters, FDEs do not escape the parameter identifiability challenge that is inherent to epidemic modeling. Like most complex dynamical systems, the fractional extensions of ODE models remain only partially observable, and depending on the choice of the fractional derivative (Caputo vs Riemann-Liouville), the initial condition identification is almost impossible. This issue is further compounded by the poor quality and reliability of epidemiological data, particularly at the onset of an outbreak, which makes outbreak modeling a compelling case study for the identification methods developed here.

### 1.1 Focus on epidemic outbreak modeling and the specificities of incidence data

Infectious diseases are one the leading causes of death worldwide. With globalization and global warming extending the endemic range of disease vectors (such as mosquitoes or ticks) into previously unaffected regions, a localized small-scale outbreak can rapidly escalate into a pandemic, as the COVID-19 demonstrated. To mitigate such extreme events, and in addition to developing pharmacological treatments, control strategies must be deployed as soon as an infectious threat is identified. These methods include: sanitary barrier, vaccination campaign and pest control for arboviruses. When preventive measures prove insufficient, broader social interventions are enforced and available treatments are mobilised at scale. Most importantly, continuous and systematic epidemic surveillance systems are now established in many countries, enabling early detection and real-time tracking of outbreak dynamics [4, 13]. Among them, mathematical models have been developed to integrate early prevalence data, predict future epidemics and identify relevant control strategies [30] but the inherent complexity of disease transmission, driven by heterogeneous host populations, environmental variability, and evolving pathogen behavior, enhances the difficulty of epidemic modeling.

Epidemic models are mostly based on the well-known SIR model introduced by Kermack and McKendrick [12] in 1927. The original population model includes three compartments: Susceptible, Infected and Recovered, a structure that can be further refined to capture pathogen specificities. Within the large existing literature on SIR models, one can cite the incorporation of seasonal effects [14, 46], the use of partial differential equations [33, 10] to account for spatial heterogeneity, or model coupling to capture cross-species transmission dynamics between animal vectors and human populations [38]. However, most of these continuous epidemic models are integer order. While a rich set of analytical tools and robust numerical schemes are available to study and simulate such systems, they often fail to capture key epidemiological features such as latency periods, multi-scale transmission dynamics, or memory effects arising from behavioral responses to an outbreak. To account for such characteristics, several alternative formalisms have been proposed, including delay differential equations [51, 43], integro-differential systems [18], or the introduction of additional compartments to enrich the model structure. Notably, fractional differential equations have established themselves as a natural and widely adopted framework in epidemic modeling [36, 37, 5], for human viral infections such as HIV [55, 57] or waterborne diseases [15], but also for animal epidemics [19] or even social media addiction [50]. Primarily because fractional derivatives encode memory and hereditary properties intrinsically, without introducing a large number of additional parameters to be estimated from data.

Yet, identifying these parameters remains complicated. Despite advances in monitoring infrastructure and treatment efficacy, predicting the severity of an epidemic in its early stages and anticipating how effective the measures taken to face it will be remains a fundamental challenge [42]. Incidence data (the number of newly reported infections) are notoriously noisy and incomplete in the early stages of an epidemic, for several compounding reasons [16]. A significant proportion of outbreaks occur in countries where systematic surveillance infrastructure is difficult to implement. Even in settings with robust monitoring policies, there is an unavoidable lag between the true beginning of an epidemic and the moment hospitals and physicians begin reporting cases, making the early phase of an outbreak practically unobservable. Furthermore, only patients who seek medical attention are recorded, while infectious individuals may transmit the disease long before symptoms manifest. These cumulative delays and reporting biases have a substantial impact on the quality of the data available for model calibration. Yet, these limitations are rarely accounted for in existing parameter estimation algorithms for FDE epidemic models, which represents a critical gap in the field.

To address this challenge, we propose a novel calibration algorithm designed to handle both partial and global observability constraints in the parameter identification of FDE epidemic models. We establish a rigorous mathematical framework for the identifiability problem in FDE systems modeling disease outbreaks, and formalise the identification problem with respect to the system structure, the fractional derivative order, and the available data. The proposed algorithm explicitly accounts for data quality, with particular attention to the unreliable observations characteristic of the early outbreak phase. It enforces the positivity of solutions throughout the numerical simulation scheme. Finally, we also investigate the minimum amount of data required for the model to reliably predict key epidemic features, such as the epidemic peak.

### 1.2 Application of the identification procedure

We apply our method on the modeling of Dengue propagation. Dengue is a viral tropical disease characterized by the flu-like symptoms such as fever, headache, or joint pain, transmitted by the bite of a female mosquito. Dengue fever affects more than 50 million people every year and it is considered by the Pasteur Institute [26] as the major human arbovirus in the world, with 55% of the global population exposed. The geographical spread of the virus keeps growing each year due to the rise of international transport and global warming. To date, there is no treatment for Dengue itself. The only way to stem the spread of the disease is to control its vector: the mosquito population and its interactions with humans. In this context, it is crucial to understand and to model how the virus behaves in a given population.

We chose Dengue as an illustrative example precisely because it concentrates multiple modeling challenges that are intrinsically tied to its dynamics. First, the mosquito life cycle introduces a complex biological layer: humans and mosquitoes operate on very different timescales, and the interactions between the two populations are difficult to disentangle. Second, like all arboviruses, Dengue exhibits a latency period between infection and symptom appearance, meaning that observed cases always lag behind actual transmission events. Third, from an identification standpoint, only the human infected population is partially observable — mosquito behavior and population dynamics are in practice impossible to record, leaving a critical part of the transmission chain unobserved. Finally, Dengue is not endemic everywhere, which means that new outbreaks can emerge in unexpected regions where surveillance infrastructure is limited or absent, further delaying data acquisition and degrading its quality in the early stages of the outbreak. Together, these challenges make Dengue an ideal and demanding test case for our identification framework.

In section 3, we present the different modeling approaches that tackle these challenges, mostly focusing on modeling the complex dynamics at play and discarding the calibration challenge behind. Starting from the classical model proposed by H. Nishiura in [38] and its generalization to the fractional case initiated by S. Pooseh and al. in [49], we then derive a new fractional model for the propagation of Dengue fever. The identification procedure is then used to define the fractional exponents as well as different parameters of the systems. The numerical simulations show that our model improves on the model proposed formerly by K. Diethelm [17].

### 1.3 Paper organization

In Section 2, we give a mathematical formulation of the identification problem as an optimization problem for a cost function over a given set of data. We then provide a numerical approach for this problem.

Section 3 applies the previous identification method to a new fractional model for the propagation of the Dengue virus. The implementation of our identification algorithm has been carried out in the programming language `R`. The construction of the identification procedure is based on two gradient-free optimization algorithms built into the environment of `R`. Our simulations show that the resulting model is more efficient than previous approaches to predict the epidemic peak in time and strength.

## 2 Numerical identification method for fractional differential equations

In this article, we focus on the identification problem for fractional differential systems. Of particular interest is the calibration of fractional models for the propagation of Dengue disease. More precisely, let ℳ_*α,π*_ be a class of fractional differential equations in the Caputo sense, i.e., fractional differential equations of the form

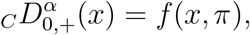

where *x* ∈ ℝ^*d*^, *α* ∈ [0, 1[^*d*^ and *π* ∈ ℝ^*m*^ is a set of parameters and

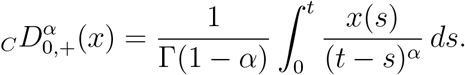

We assume that we have observations of a dynamical process over a given time scale denoted by 𝕋_*s*_ on a given interval [0, *T*], where *s* ∈ ℕ, and given by

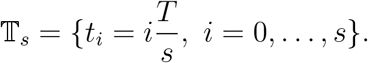

The observations are defined by the data of a function 𝒪: 𝕋_*s*_ →ℝ^*k*^, *k* ≤ *d*, which is known. When *k* = *d* (resp. < *d*) we have the case of *global observations* (resp. *partial observations*).

We assume that the dynamical process under consideration corresponds to the model associated to 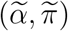 in the class ℳ _*α,π*_.

One of the major problems is then:

- Can we recover the value of 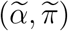 from the known observations 𝒪 on 𝕋_*s*_?

To answer this question, we formalize this problem as an optimization problem. It can be proved under weak assumptions that such a minimization problem admits minimizers. Of course, the main problem is to compute the minimizers. This can be done using a specific numerical procedure that we describe in this paper. The regularity of the underlying functional is no more than a continuous inducing difficulty in the numerical optimization process. In particular, we have to avoid gradient-dependent numerical methods.

The identification of the model depends on the time scale 𝕋_*s*_ as well as the quality of the data. A set of natural questions in this setting is as follows:

- How small can be 𝕋_*s*_ in order to have a good approximation of 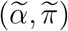 ?
- How the quality of data over a fixed 𝕋_*s*_ impact the identification of 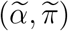 ?

These two questions are crucial when applying an identification method to the propagation of epidemics. The first one relies on the efficiency of identification for prediction purposes during the early stage of the epidemic. The second one deals with the classical problem of aggregating data for a given epidemic in the early and late stages of a given outbreak.

From a practical point of view, in many applications, we are not interested by the precise long-time behavior of the system but more in specific quantities which are relevant from the prediction point. For example, in an epidemic problem, we are interested in obtaining information as soon as possible concerning the amount of data, the possibility of predicting the time and strength of the epidemic peak with sufficient precision. By sufficient precision, we understand that the obtained prediction for this peak will not be drastically changed by new data.

To answer all these questions, we need to construct an efficient numerical identification method for fractional dynamical systems. From the theoretical point of view, this is a classical approach to the optimization problem adapted to fractional differential equations. The efficiency of the optimization process will depend on the quality of the discretization process of the solutions of the fractional equation.

### 2.1 Identification problem

Let *α* ∈]0, 1]^*d*^, *K* a compact subset of ℝ^*m*^ and *f* : (*x, π*) ∈ ℝ^*d*^ × *K* →ℝ^*d*^ a function which is Lipschitz with respect to *x* and continuous with respect to *π* ∈ *K*, where *π* stands for a set of model parameters which can be identified.

We assume that *x* can be modeled by an element *M*_*α,π*_ of class ℳ_*α,π*_. We denote by ℒ (*α, π*) the cost-function defined as the root relative squared error (RRSE) of the form

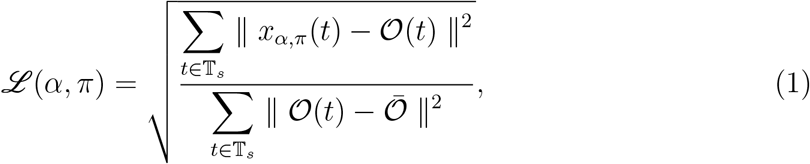

where *x*_*α,π*_ is the solution of the model *M*_*α,π*_ such that *x*_*α,π*_(*t*_0_) = 𝒪 (*t*_0_), and 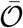 is the average of the given observations. The proposed error is relative to the situation of using the simple predictor as the average of the real data. The values of ℒ are in the range [0,∞ ), where 0 means the perfect fitting, i.e. *x*_*α,π*_(*t*) = 𝒪 (*t*), for all *t* ∈ 𝕋_*s*_. When the cost-function ℒ exceeds the value of 1, it means that obtained predictor is worse than the simplest predictor in the form of average.

The previous functional is well defined as we have existence and uniqueness for each (*α, π*) of the solution *x*_*α,π*_ due to the Lipschitz assumption on *f* with respect to *x* (see [17], Theorem 6.5 p.93).

The fractional identification problem corresponds to the optimization problem :

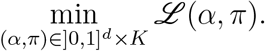

It must be noted that the functional ℒ (*α, π*) depends continuously on *α* and *π*. This is a consequence of the two following results:

- the continuous dependence of the solutions of a fractional differential equation with respect to *α* in one hand (see [17], Corollary 6.23 p.216);
- the continuous dependence of the solutions with respect to the parameters *π* in a second hand (see [17], Theorem 6.21 p.215).

As we look for the minimization of ℒ (*α, π*) on a compact set]0, 1]^*d*^ × *K*, there exists at least one minimizer (*α*_⋆_, *π*_⋆_) by the classical Weierstrass argument.

Any minimizer (*α*_⋆_, *π*_⋆_) of the optimization problem leads to a model 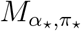 which can be considered as an efficient description of the dynamical process.

In practice, minimizers are obtained using numerical algorithms. A proposed algorithm is described in the next section.

### 2.2 Numerical approach to the optimization problem

To implement optimization issue, we need to solve numerically *M*_*α,π*_. From the theoretical point of view, we can adapt any convergent and stable numerical scheme for solving fractional differential systems. Once we obtain a numerical solution to *M*_*α,π*_, we can proceed with the optimization algorithm. We propose the procedure of finding an approximate minimizer of the cost function (1), which is presented in the Figure 1.

**Figure 1.**
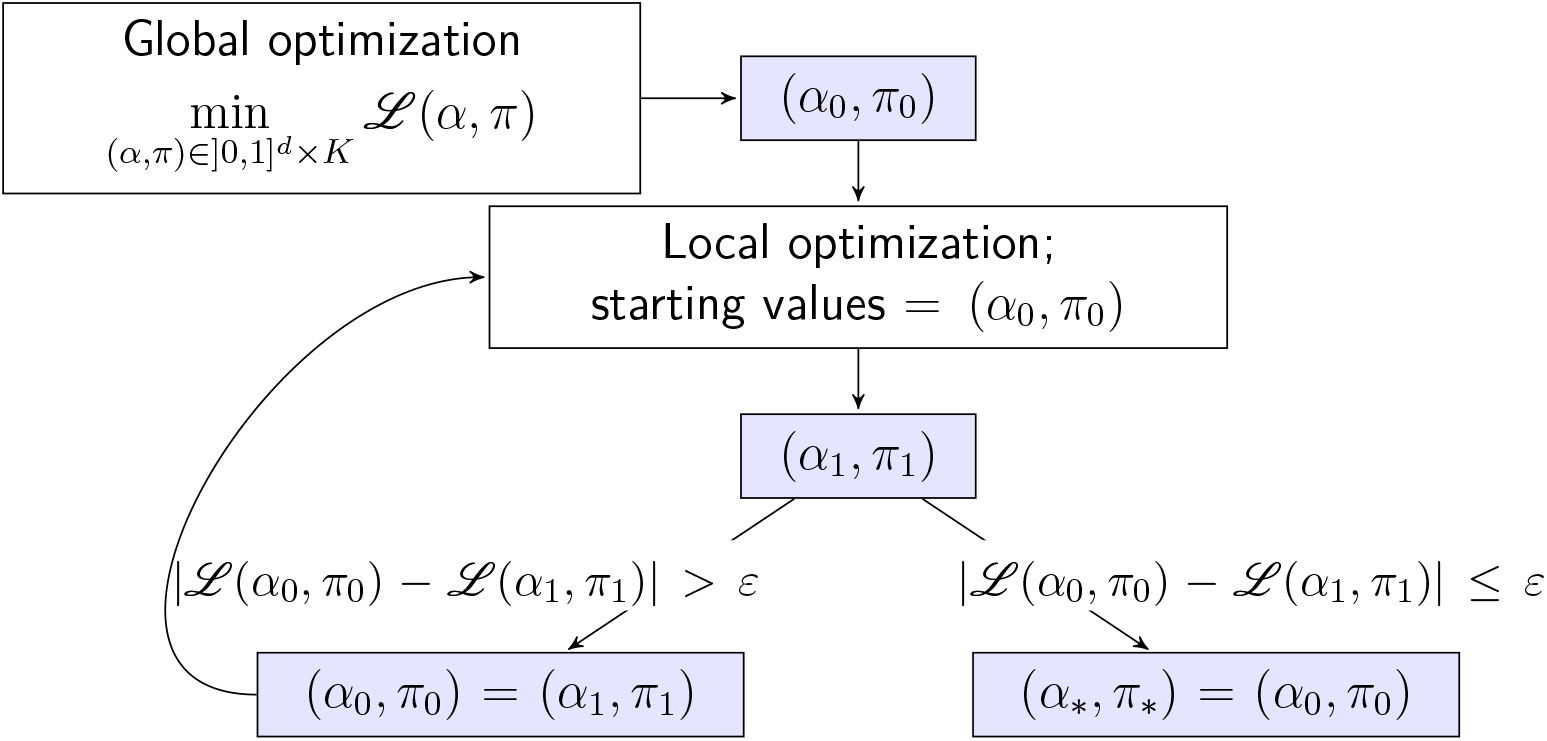
Scheme of the optimization algorithm

**Figure 2.**
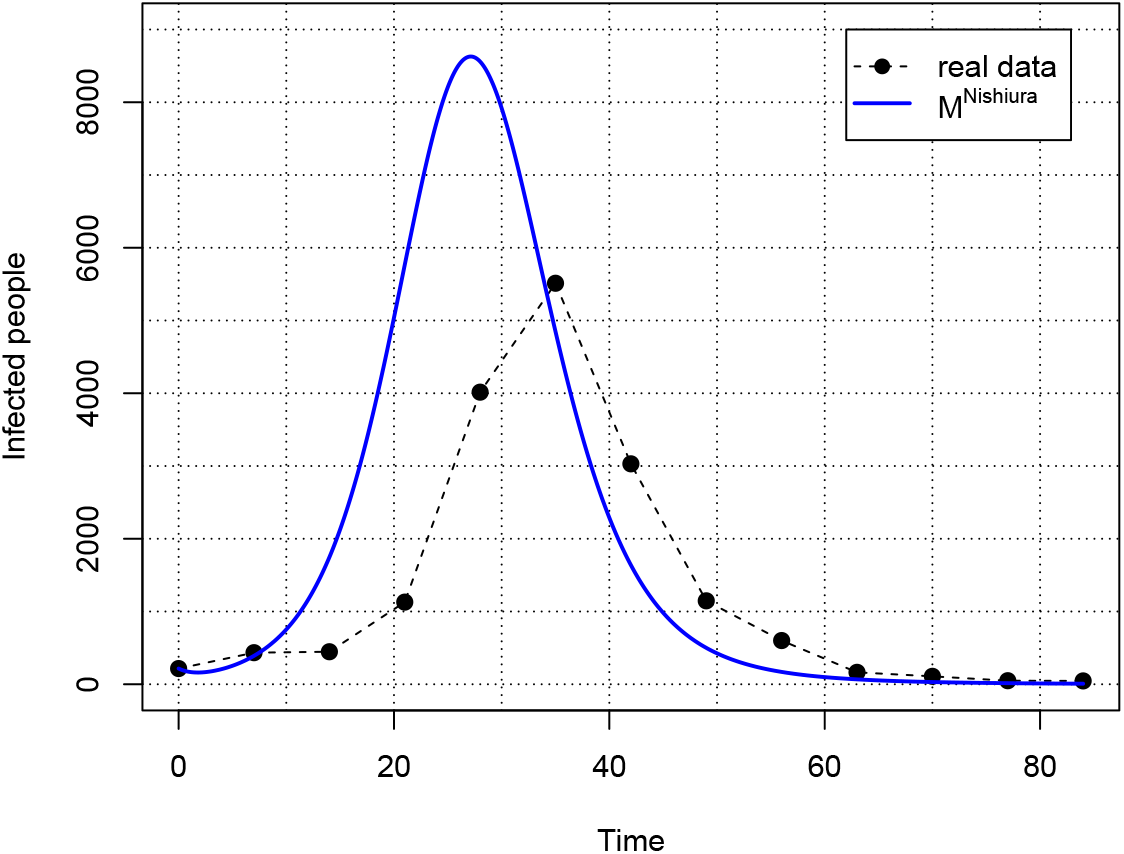
Nishiura classical SIRSI model

In most cases, it is difficult to observe if the cost function (1) possesses only the global optimum or many local optima. As we are interested in finding the approximate global optima, we propose to start with the global optimization algorithm to catch the area of possible global minimizers. The global optimization algorithms do not require a starting point, which is chosen randomly. As a consequence, the convergence to the global minimizer can run very slowly. The idea is to use the global optimization algorithm to find the initial point for the local optimization procedure, which, with the appropriate algorithm, can be fast convergent. We repeat the local optimization procedure with the starting point coming from the previous one until the change of the value of the cost function does not improve results significantly.

### 2.3 Positivity-preserving schemes for fractional population models in Biology

Our applications deal with a class of fractional population dynamical models introduced in ([27],p.429 and Theorem 10 p.430). These models satisfy a fundamental property called positivity: when *x*_0_ ≥ 0, then *x*(*t*) ≥ 0 for all *t* > 0. We denote these class by 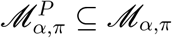 . They are of the form:

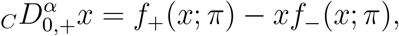

where *f*_+_, *f*_−_ are positive with *x* ≥ 0 and Lipschitz continuous.

The previous numerical approach to the optimization problem assumes that an efficient numerical scheme is provided. For systems in 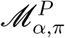, a numerical scheme must then preserve positivity. Otherwise, we can obtain simulation results that are not possible to interpret. This problem was studied in [27] where a **nonstandard numerical scheme** is constructed. This numerical scheme will be used in our application of the identification procedure to the fractional homogeneous Nishiura model for the Dengue propagation in Section 3.

### 2.4 Accounting for unreliable data at the margins of an outbreak with a weighted cost

In most cases, the available observation data of a given epidemic have two kinds of quality. At the beginning of the epidemics, not all people are tested, or institutions attributes some symptoms to another disease, leading to poor quality data. In particular, we expect an underestimated number of infected people. When the epidemic is well established, all actors for disease control are efficient and produce more valuable data. In the same way, when the outbreak is finishing, the control measures are less stringent inducing poor quality data.

To take account this variable quality of data, we introduce a weight function *w*(*a*) : 𝕋_*s*_ → ℝ^+^, where *a* is the weight hyperparameter to identify. This weight function helps distinguish between early and late stage epidemic data and the main evolution time of the outbreak. Our cost function therefore 1 becomes:

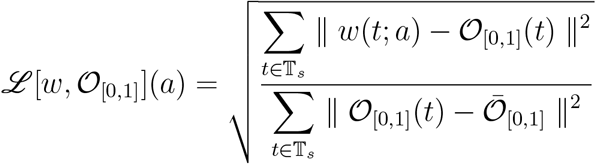

An example of such a weight function applied to epidemic data is constructed in Section 3.4.1. The underlying principle is general: time points where observations are reliable and noise is low are assigned large weights, while time points affected by acquisition uncertainty, underreporting, or high noise levels are downweighted, reducing their influence on the calibration.

In order to illustrate the previous numerical method of identification, we provide in the following section an application to the modeling of the Dengue fever propagation. After an introduction to Dengue modeling approaches, we introduce a new fractional model called fractional homogeneous Nishiura model based on previous work of S. Pooseh and al. [49], K. Diethelm [17] and T. Sardar and al. [53]. Using a data set of the 2009 Dengue outbreak in Cape Verde islands, we give the result of the weighted and non weighted identification procedure. In both cases, we observe that we have a fractional dynamics close to a classical one for humans but not for mosquitoes.

## 3 Applications results: Modeling Dengue fever outbreak

Dengue is a tropical viral disease transmitted by female Aedes mosquitoes (Ae. aegypti, Ae. albopictus, Ae. polynesiensis), affecting over 50 million people annually across more than 100 endemic countries in Africa, the Americas, Asia, and across Pacific Ocean, with the Institut Pasteur identifying it as the most significant human arboviral disease, exposing over 55% of the world’s population and representing a leading cause of hospitalisation and mortality, particularly among children and adults in Asia and Latin America. The disease presents in four serotypes (DEN-1 to DEN-4) with no cross-immunity, manifesting as influenza-like symptoms — fever, joint pain, headache and nausea — but capable of progressing to severe haemorrhagic Dengue, which is frequently fatal within 48 hours and is notably more likely upon reinfection with a heterologous serotype.

After biting an infectious human — who remains contagious for 4 to 12 days during illness — the female mosquito undergoes an extrinsic incubation period of 4 to 10 days before becoming capable of transmitting the virus (we will hereafter refer to mosquitoes exclusively as females, since only females bite humans). Once infected, the mosquito remains contagious for the remainder of its life. Thriving in warm, humid environments with standing water that favours reproduction, the vector shows a marked preference for urban and peri-urban areas of tropical and subtropical countries.

During its development, the mosquito passes through three stages before reaching adulthood: the female first lays her eggs in water, which subsequently hatch into larvae before undergoing pupation. Once adult, its lifespan is approximately 50 days. Notably, the primary driver of disease spread remains the human host, as the mosquito spends its entire life within a restricted perimeter close to its breeding site, rarely travelling more than 400 to 500 metres.

The mosquito feeds during the day, with biting activity peaking in the early morning and at dusk. Finally, the mosquito exhibits a remarkable memory effect when locating potential hosts: if a female identifies a site where she has reliable access to a sufficient number of humans, she will tend to return to that location on subsequent days.

### 3.1 Classic modeling approaches for Dengue outbreaks

The majority of existing models of Dengue outbreak incorporate both human and mosquito dynamics. But in practice, it is impossible to take into account all the characteristics of a Dengue outbreak, such as climate impact, vector behavior or demographic and sociodemographic specificities, in one unique model with a reasonable number of parameters. Therefore, models usually focuses on one specific feature of Dengue outbreaks [41, 39, 31, 6, 44].

One of the most established trends in Dengue modeling focuses on the interaction between multiple circulating serotypes and the role of temporary cross-immunity [6, 44]. They typically use SIR or SEIR structures, subdivided by serotype and infection history which can be used to mimic the impact of vaccination policies for instance [28]. Recent models bridge the gap between individual-level immune responses and population-level transmission dynamics. Coupled Multiscale Models are used, combining within-host ODEs (tracking viral load and antibody levels) with population-level Partial Differential Equations (PDEs) or structured transmission models. This allows researchers to track host antibody distributions across the population.

Another important trend is climate-induced changes [31]. Meteorological indicators are used to trigger alarm indicators announcing future Dengue outbreaks with temperature and rainfall the most frequently used predictors. Several models also take into account the seasonal variation of mosquitoes behaviors.

Finally, from a methodological point-of-view, the majority of existing models are deterministic and continuous systems but other formalisms must be mentionned. Discrete frameworks such as Agent-based Models and stochastic equations have also been designed to study demographic stochasticity and spatial heterogeneity, and data-driven methods based on machine-learning are prevalent now in the literature [31]. Last but not least, in the absence of treatment and effective vaccines, vector control strategies targeting mosquitoes have been extensively modeled [39]. They include the release of sterile insects or the use of chemical compounds.

To illustrate our method here, we chose one of the simplest existing ODE model of Dengue outbreak, the model of Nishiura.

#### 3.1.1 Focus on the classical Nishiura’s model

In [38], H. Nishura defined a new compartmental model for which he explores the seasonality and the periodicity of the disease, the parameters meaning and values for special outbreaks. In particular, he computes the reproduction rate.

The model couples an SIR system for the humans and a SI for the mosquitoes, which are believed to never recovered after being infected. As a consequence, the human population (host) is splitted in three compartments: *S*_*h*_, *I*_*h*_ and *R*_*h*_ (susceptible, infected and recovered) and only two classes for mosquitoes, *S*_*m*_ and *I*_*m*_. The Nishiura model is given by:

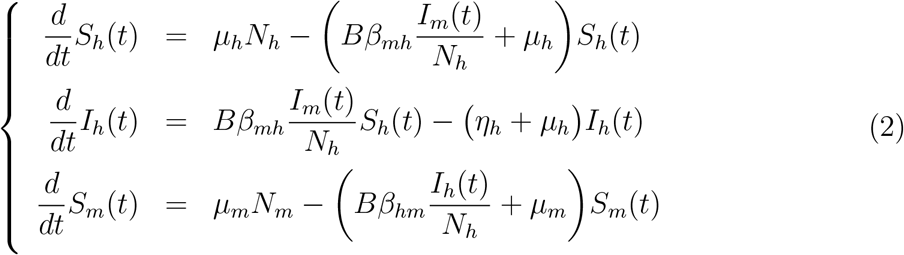

where *R*_*h*_ = *N*_*h*_ − *S*_*h*_ − *I*_*h*_ and *I*_*m*_ = *N*_*m*_ − *S*_*m*_, with the parameters:

*µ*_*h*_ (respectively *µ*_*m*_) - the natural death rate for humans (resp. for mosquitoes);

*η*_*h*_ - the host recovery rate;

*B* - the average of bites per day;

*β*_*mh*_ (resp. *β*_*hm*_) - the related rate to the transmission probability per bite from infected mosquito to human (resp. infected human to mosquito).

The assumption to have a constant human population is due to the fact that the period of observation is short. For the mosquito population, as no specific actions are made in order to drastically reduce the population, we can assume also that it is constant on the given period.

The model parameters have been already identified by H. Nishiura in [38] using Dengue outbreaks in Brazil. These parameters can be divided into two groups:

- One dealing with human population in the specific area of observation.
- One dealing with behavior of the mosquito population under study.

For the human population parameters, they are obtained statistically over the population and are provided by governmental institution. They change according to place.

The mosquito parameters are of biological origin and depend mainly on the type of mosquitoes under study. As the mosquito species are the same in Cape Verde and Brazil, these parameters remain the same as in the Nishiura’s model.

The simulations of (2) are done using the numerical method mentioned in Section 2. We take as initial conditions the values known from government statistics, i.e.

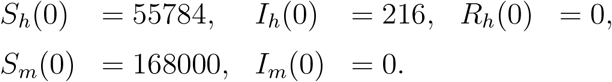

We see that the classical Nishiura model is not satisfying for predicting the peak of epidemics in term of time or strength. As the parameters in the system (2) are model independent, they can not be considered as identifiable parameters. As a consequence, there is no possibility to improve the model results by any identification procedure.

Therefore the model must be improved in itself. Without entering in the complexity of the Dengue disease and its propagation, we can look for a model taking in consideration one important aspect of the mosquito behavior. Actually, if the female mosquito discovers a place where it is easy to get enough people to bite, will come back the day after. For more details, see [40, 26, 23, 32, 58] [40] - [32]. As a consequence, we must take into account some memory effect of the dynamics of mosquitoes.

A classical approach to deal with memory effects is to use fractional dynamics. The next two sections therefore introduces fractional formalism to account for mosquitoes preferences. Section 3.2 describes the existing fractional models that have be implemented to model mosquitoes preferences but also their drawbacks while in Section 3.4, we introduce a fractional version of Nishiura that preserves all the advantages of Nishiura model while also accounting for mosquito memory.

### 3.2 Previous Fractional Dengue fever epidemic models

Fractional models have emerged as a significant advancement in understanding and predicting Dengue outbreaks and therefore have been widely applied recently. These models use derivatives of non-integer order to capture the complex dynamics of Dengue transmission more accurately than traditional integer-order models [47], fitting real-world data more closely, as demonstrated in studies conducted in regions like Lahore, Pakistan [9], Selangor, Malaysia [35] or in Nepal [22]. The fractional approach allows for a more nuanced representation of the disease’s spread, accommodating the memory and hereditary properties of the transmission process. This is particularly important in modeling infectious diseases like Dengue, where the interaction between human and mosquito populations is intricate and influenced by various factors.

These models use more or less complex SIR-like models, focusing on model’s calibration, reproduction score with parameters sensitivity [34, 45, 21] or implementing control strategies [54, 1, 3] through mosquito population control [48] or vaccination [20, 45, 56]. But they do not implement any strategy to preserve numerical solution positivity or adapt their calibration strategy to the fact that outbreak data are more reliable closer to epidemic peak than at the early beginning of the epidemic.

The first fractional model for the Dengue epidemics was proposed by S. Pooseh and al. in [49] obtained by replacing directly the classical derivative in the Nishiura model by the Riemann-Liouville fractional derivatives. However, as already noticed by K. Diethelm ([17],§.2.2 p.616) such models have several drawbacks:

- The classical assumption about the constancy of the total population for human and mosquitoes over the period of interest is broken.
- Units of the biological parameters of the models must be changed because the left hand side is in *t*^−*α*^ instead of *t*^−1^. As a consequence, the meaning of the biological constant has to be changed.
- Riemann-Liouville differential equations lead to singular solutions when fixing initial conditions. As a consequence, the model is not well posed.
- The fractional dynamical parameter is assumed to be the same for humans and mosquitoes, which has no biological support.

Part of these problems were solved in the fractional models proposed by K. Diethelm in [17]. He considers the following fractional models:

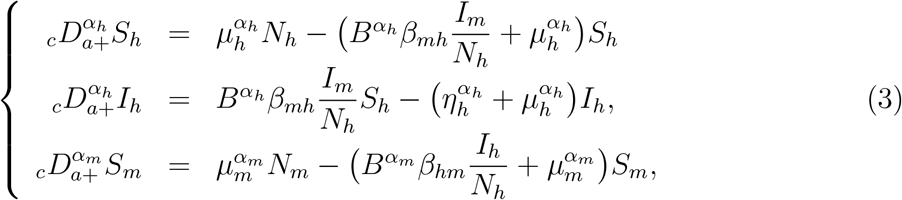

with *R*_*h*_ = *N*_*h*_ −*S*_*h*_ −*I*_*h*_, *I*_*m*_ = *N*_*m*_ −*S*_*m*_ and *α*_*h*_ ∈]0, 1], *α*_*m*_] ∈ 0, 1] which is different from ours.

The use of Caputo fractional derivative avoid many problems related to the initial conditions problems as well as the well-posedness of the problem and the assumption of the constancy of the populations. Indeed, the Caputo derivative is defined as a fractional integral of an integer-order derivative, which means its initial conditions take the same form as those of classical ODEs. These are physically interpretable and measurable quantities. By contrast, the Riemann-Liouville fractional derivative requires initial conditions that are fractional-order derivatives of the solution at *t* = 0 that have no clear physical meaning and are essentially impossible to measure or estimate from data, especially in epidemiology. In addition, because the Caputo derivative preserves the structure of classical differential operators at the initial time, the standard functional analysis tools carry over with relatively minor adaptations. This makes it far easier to establish existence and uniqueness theorems.

The homogeneity of the fractional differential system (3) is also satisfied by taking on the right-hand side the biological parameters to the power of an appropriate alpha. However, as noticed by A. Dokoumentzidis and al. in [2] and by T. Sardar and al. in ([53], note p.513), as different powers of some parameters are used (as for *B* for example), the significance of these new parameters is not clear.

A new fractional model is then proposed by T. Sardar and al. in [53] following a strategy developed in [2]. However, such a model leads to several questions:

- As already noticed in ([53], p.514), the obtained fractional model ((2.4), p.514) is a priori ill-defined due to the use of the Riemann-Liouville fractional derivatives. The replacement of the Riemann-Liouville derivative by the Caputo one is only valid for the asymptotic of the solutions, meaning when *t* goes to infinity. However, for what concerns the prediction of the evolution of the Dengue disease, we are interested mainly in finite time dynamics, as for *t* going to infinity, most of the time the epidemics disappears for a sufficiently long time.
- To deal with classical initial conditions, the authors propose to use the classical relation between the Caputo and the Riemann-Liouville fractional derivative. Then, the initial model ([53],(2.4)) is replaced by another one where Caputo derivatives are used, thanks to the introduction of correction terms depending on the initial conditions and supporting the singularities. As a consequence, we do not have a model in the classical sense, but an infinite family of models depending on initial conditions. The problem is then to interpret this model dependence from a biological viewpoint. Indeed, a model is intended to be derived from laws which are a priori universal in time and space and precisely not dependent on your initial time of modeling or initial point of departure.

In the following, we propose a new fractional model using fractional homogeneity to solve the previous difficulties. More precisely, we generalize the classical Dengue fever epidemic model defined by Nishiura to a fractional one following previous work of S. Pooseh and al. [49], K. Diethelm [17] and T. Sardar and al. [53].

### 3.3 Dengue fever outbreak in 2009 in Cape Verde

To allow a fair comparison, we consider the same data set from [49]. They consist in the number of new infected people for the last 12 weeks of 2009 for the Dengue fever outbreak in Cape Verde:

We refer to [58] for a better understanding of the amplitude of this outbreak. As only the number of new infected is available, we are here in the situation of a partial observable system.

### 3.4 A new fractional homogeneous Nishiura model

To deal with restoring **homogeneity of fractional embedded equations** we use one of the methods described in P. Inizan [24] (see also [25]).

The dimension of classical (differential) time operator is *T*^−1^. We focus on the construction of a fractional operator in our mathematical model, whose dimension is the same as the classical one. For this purpose, we introduce a *d*-dimensional constant quantity *τ* ∈ ℝ^*d*^ homogeneous to time, i.e. of dimension homogeneous to *T* . Multiplying directly the left side of the fractional equation (2) by *τ*^*α*−1^, we obtain on the left side a fractional operator with dimension *T*^−1^. Since the time constant *τ* is defined independently of the classical equation it needs to be taken into consideration in the identification problem.

From the dynamical viewpoint, the previous modification of the fractional derivative can be understood as follows:

Let *τ* > 0, we denote by ⋆_*τ*_ the operator defined on functions *x* ∈ *C*([0, *T*]; ℝ^*d*^) by

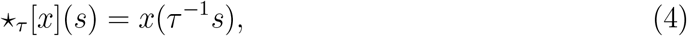

for *s* ∈ [0, *τ T*]. We have:

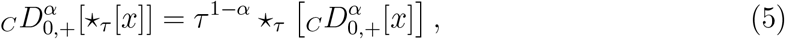

which corresponds to the following commutative diagram:

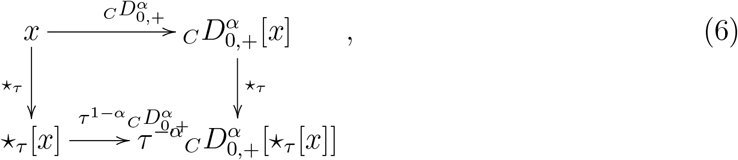

Indeed, we have

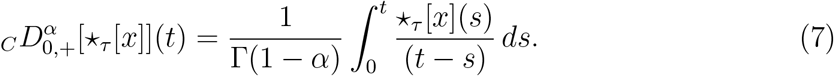

Making the change of variables, *u* = *s/τ*, we obtain

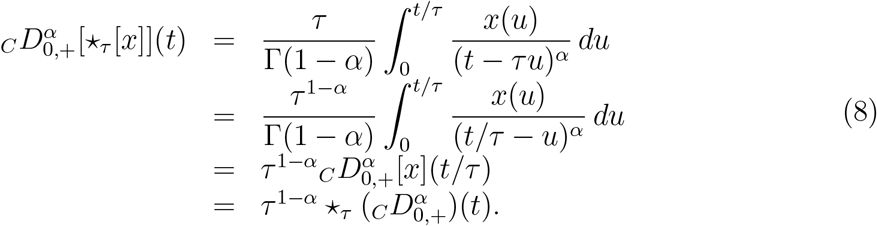

Considering two exponents *α*_1_, *α*_2_ and two variables *x*_1_, *x*_2_ satisfying the fractional differential system

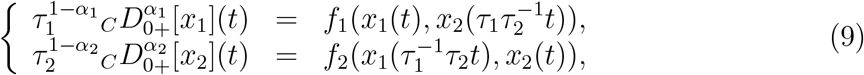

that the image of a solution (*x*_1_(*t*), *x*_2_(*t*)) by the operators 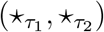 is a solution of the following fractional homogeneous differential system

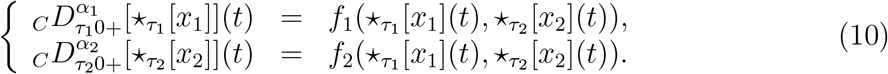

As a consequence, we see that multiplying by some constant time the fractional derivative is equivalent to take into account the different scale of dynamics for the processes under consideration. In particular, the scale 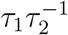 plays a fundamental role.

For the modeling of the Dengue disease we have two different populations: human and mosquitoes. As a consequence, we wait for two distinct time parameters *τ*_*h*_ and *τ*_*m*_.

The procedure then leads to the following model:

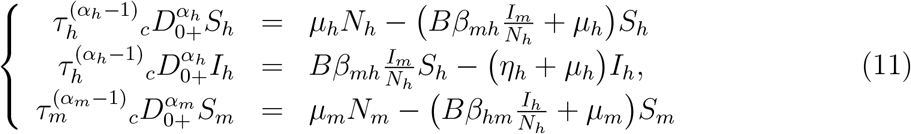

with *R*_*h*_ = *N*_*h*_ − *S*_*h*_ − *I*_*h*_, *I*_*m*_ = *N*_*m*_ − *S*_*m*_.

Moreover, as explained before, a special role is devoted to the quantity 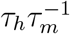. We expect that the intrinsic scale for the dynamics of mosquitoes is fast with respect to the intrinsic scale for the dynamics of humans. As a consequence, we can assume that *τ*_*m*_ ∈ [0, 1] and *τ*_*h*_ *≥*1. This remark has important consequences on the minimzation procedure. Indeed, instead of minimizing the cost function ℒ (*α*_*h*_, *α*_*m*_, *τ*_*h*_, *τ*_*m*_), we will apply our procedure to the functional 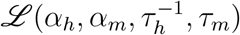 for which all the variables belong to [0, 1]^4^. As a consequence, the classical Weierstrass argument implies that there exists at least one minimizer for this functional, justifying the application of our numerical algorithm.

#### 3.4.1 Model’s calibration using 2009 Cape Verde Dengue outbreak and our numerical algorithm

As it was mentioned in Section 2.3, to solve numerically the fractional homogeneous Nishiura model (11) we adopt the nonstandard numerical scheme described and analyzed in [27].

We apply our numerical identification procedure for partially observed systems by choosing as a global optimization process the *global optimization by differential evolution* implemented in `R` by the function `DEoptim` and for the local optimization process the *Constrained Optimization BY Linear Approximation* implemented in`R` by the `nloptr` function with the algorithm `NLOPT_LN_COBYLA`. Both methods are gradient free optimization algorithms avoiding classical difficulties with the application of classical gradient like methods for optimization of fractional differential systems.

The identification procedure was developed in two cases: the classical one described in Section 2.2 and the one considering the quality of data. To enhance the impact on the identification of data from the main course of the epidemic, we introduce the weight function *w* : 𝕋_*s*_ → ℝ^+^ which is constructed according to the available data using exponential function

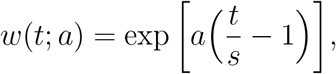

where *a* is the weight parameter to identify.

The main idea behind this weight function is that at the beginning of the epidemic, not all infected people are tested or declared as infected. However, as the epidemic develops, people are tested more systematically. A way to take this phenomenon into account is to introduce a weight whose evolution depends on the epidemic.

In order to identify the weight parameter *a* first, we sort the given set of data in ascending order. Next, we normalize the data using the min-max normalization. Let the normalized data be defined as 𝒪 _[0,1]_. Then we find the minimum of the following cost-function

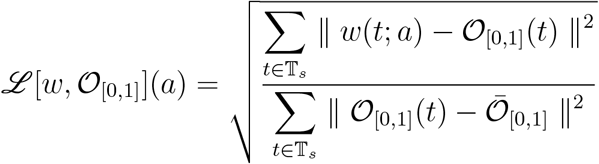

using the non-linear minimization based on the Newton-type algorithm implemented in `R` as `nlm` function.

The example of the weight function in the case of Dengue data is presented on the Figure 3.

**Figure 3.**
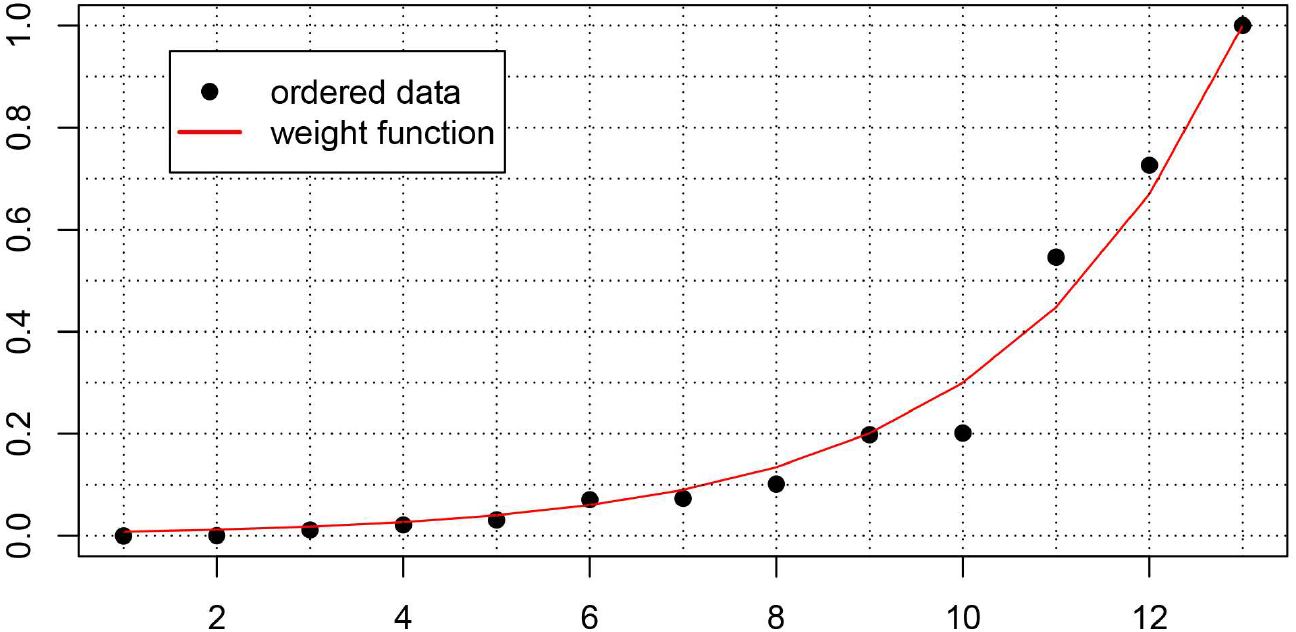
Sorted and scaled data (min-max normalization) and fitted weight function

Then we define the weighted cost-function as follows:

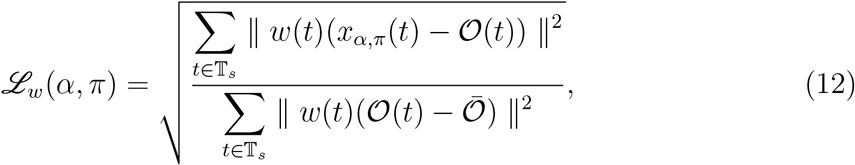

The corresponding identification algorithm implementation is available on request.

#### 3.4.2 Identification of the fractional homogeneous model

Applying the previous algorithm on the fractional homogeneous Nishiura model estimating only the parameters *α*_*h*_, *α*_*m*_, *τ*_*h*_ and *τ*_*m*_, we obtain the results presented in the Figure 4.

**Figure 4.**
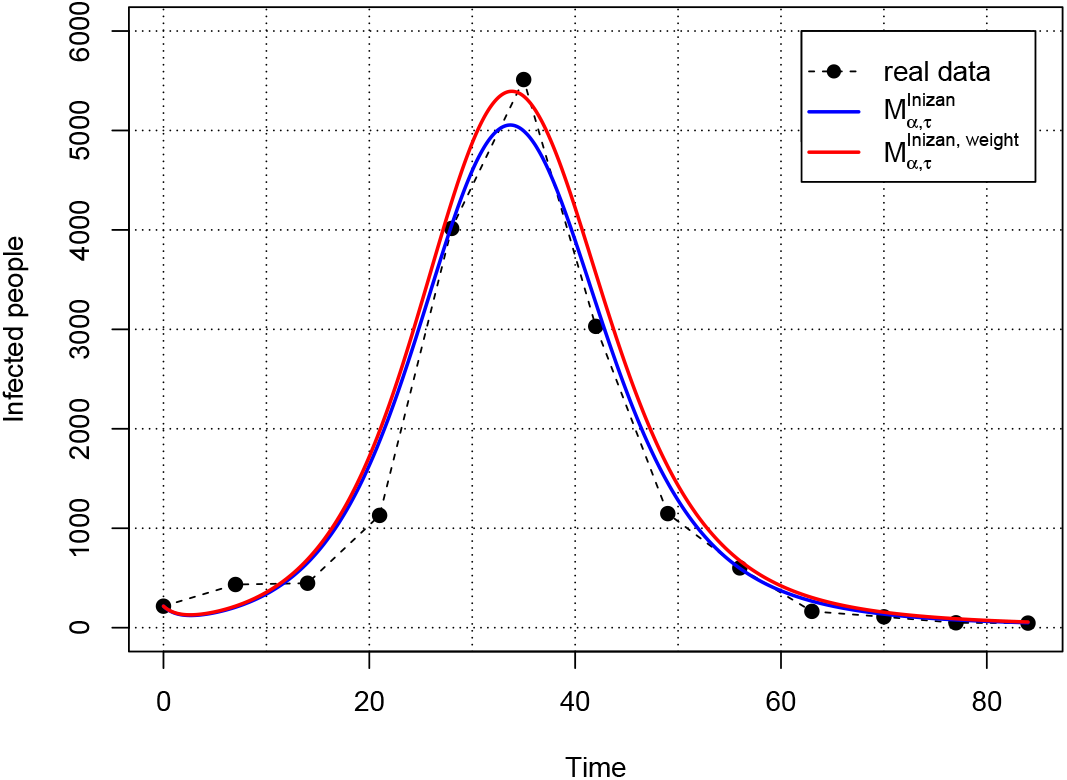
Result of the fitting procedure for the fractional Nishiura model with and without weights

We can observe a good agreement between the obtained fractional model in both cases. However, the weighted version gives a better prediction for the strength of the epidemic peak. It suggests that weighted data have to be considered for identification purposes.

Moreover, we can observe that the fractional dynamics for humans is closer to a classical dynamics then for mosquitoes. This result is coherent with the assumption that mosquitoes have a memory effect which must be taken into account for the modeling.

### 3.5 Comparison with previous fractional approaches

#### 3.5.1 Comparison with the Diethelm’s fractional model

We have applied our algorithm to the Diethelm’s fractional model for the Dengue given by (3). We have obtained the results presented on Figure 5.

**Figure 5.**
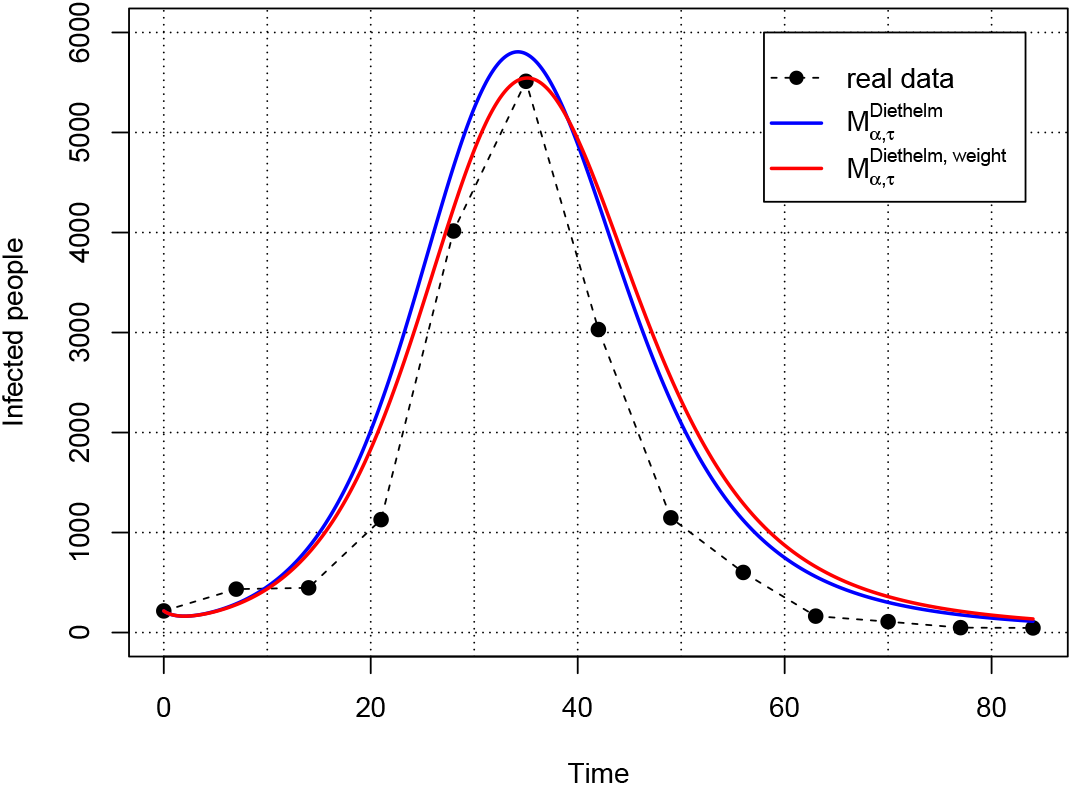
Result of the fitting procedure for the Diethelm model with and without weights

The global fitting of the data is less satisfying than with our model. In particular, the end of the epidemics is not well modeled.

In the Table 3 we collected the results of the parameters identification, with and without weights, for the model proposed by Diethelm and for the new model proposed in the paper. In both cases of identification, with and without the weight function, the RRSE value is about two times smaller for the model proposed by the authors of this paper.

**Table 1:** Parameter values in Nishiura’s model.

| Significance | Parameter | Value |
| --- | --- | --- |
| Per capita mortality of humans | $\mu_h$ | $\frac{1}{71 \times 365}$ |
| Per capita mortality of mosquitoes | $\mu_m$ | $\frac{1}{10}$ |
| Transmission probability from human to mosquito | $\beta_{hm}$ | 0.36 |
| Transmission probability from mosquito to human | $\beta_{mh}$ | 0.36 |
| Recovery rate of human | $\eta_h$ | $\frac{1}{3}$ |
| Biting rate | $B$ | 0.7 |

**Table 2:**
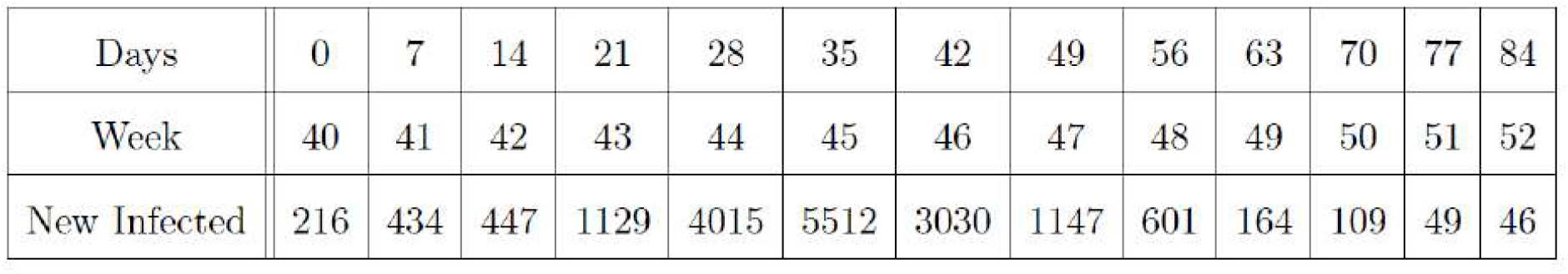
Reported incidence data for the 2009 dengue fever outbreak in Cape Verde.

|  |  |  |  |  |  |  |  |  |  |  |  |  |  |
| --- | --- | --- | --- | --- | --- | --- | --- | --- | --- | --- | --- | --- | --- |
| Days | 0 | 7 | 14 | 21 | 28 | 35 | 42 | 49 | 56 | 63 | 70 | 77 | 84 |
| Week | 40 | 41 | 42 | 43 | 44 | 45 | 46 | 47 | 48 | 49 | 50 | 51 | 52 |
| New Infected | 216 | 434 | 447 | 1129 | 4015 | 5512 | 3030 | 1147 | 601 | 164 | 109 | 49 | 46 |

**Table 3:** The numerical results: Parameter values after calibration with and without weights for Diethelm model and the Fractional Homogeneous Nishiura model.

| Model | $\alpha_h$ | $\alpha_m$ | $\tau_h^{\alpha_h-1}$ | $\tau_m^{\alpha_m-1}$ | $\mathcal{L}$ | $\mathcal{L}_w$ |
| --- | --- | --- | --- | --- | --- | --- |
| $M_\alpha^{Diethelm}$ | 1 | 0.796 | — | — | 0.393 | — |
| $M_\alpha^{Diethelm,weight}$ | 1 | 0.778 | — | — | — | 0.178 |
| $M_{\alpha,\tau}^{Inizan}$ | 0.994 | 0.957 | 0.72 | 1.619 | 0.171 | — |
| $M_{\alpha,\tau}^{Inizan,weight}$ | 0.993 | 0.944 | 0.778 | 1.540 | — | 0.088 |

#### 3.5.2 Comparison with the T. Sardar et al. fractional model

We have applied our numerical algorithm in order to calibrate the model proposed by T. Sardar et al. in [53]. For the simulations, we have used the numerical algorithm proposed in [53], implemented in `Matlab`, in order to have reliable numerical experiments with the one already provided in [53]. We refer to [52] for a complete discussion of the model and numerical algorithm proposed by T. Sardar et al. in [53].

The identification of parameters leads to the fitting result of the Cape Verde outbreak presented in Figure 6.

**Figure 6.**
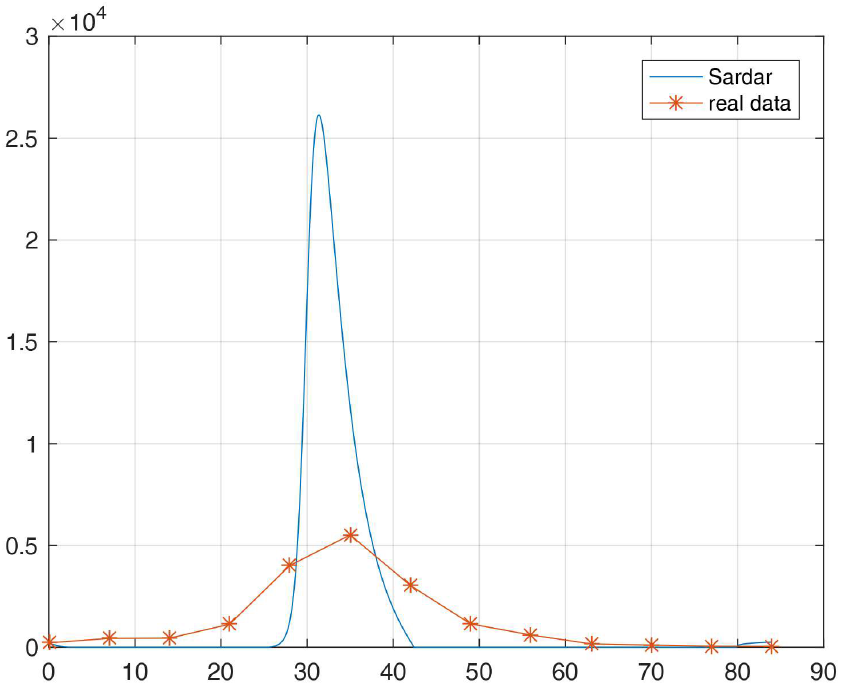
Result of the fitting procedure for the Sardar et al. model

As can be seen, the fitting is very bad. This is due in part to the use of the Riemann-Liouville fractional derivative in the modeling which induces a strong singularities at the beginning of the dynamics. The control of this singularity leads to inadequate values for the fractional parameters.

### 3.6 Epidemic outbreak predictions for a short term observation - efficiency of identification

Assume now that we would like to know how many information is needed, i.e. how long we need to observe the epidemic, to predict the strength and time of the epidemic outbreak.

The natural question arises: what is the smallest subset **T**_*k*_ of 𝕋_*s*_ (the smallest *k* ≤ *s*)

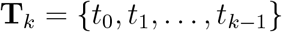

where *k* = 1, …, *s*, such that 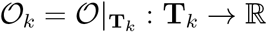 gives a good prediction of 𝒪?

To answer this question we use the identification algorithm described in the article for different length of data set. The results of the predictions obtained with Nishiura’s fractional model we compare with predictions by Diethelm’s fractional model in both cases, without and with the weights (Figure 7 and Figure 8, respectively).

**Figure 7.**
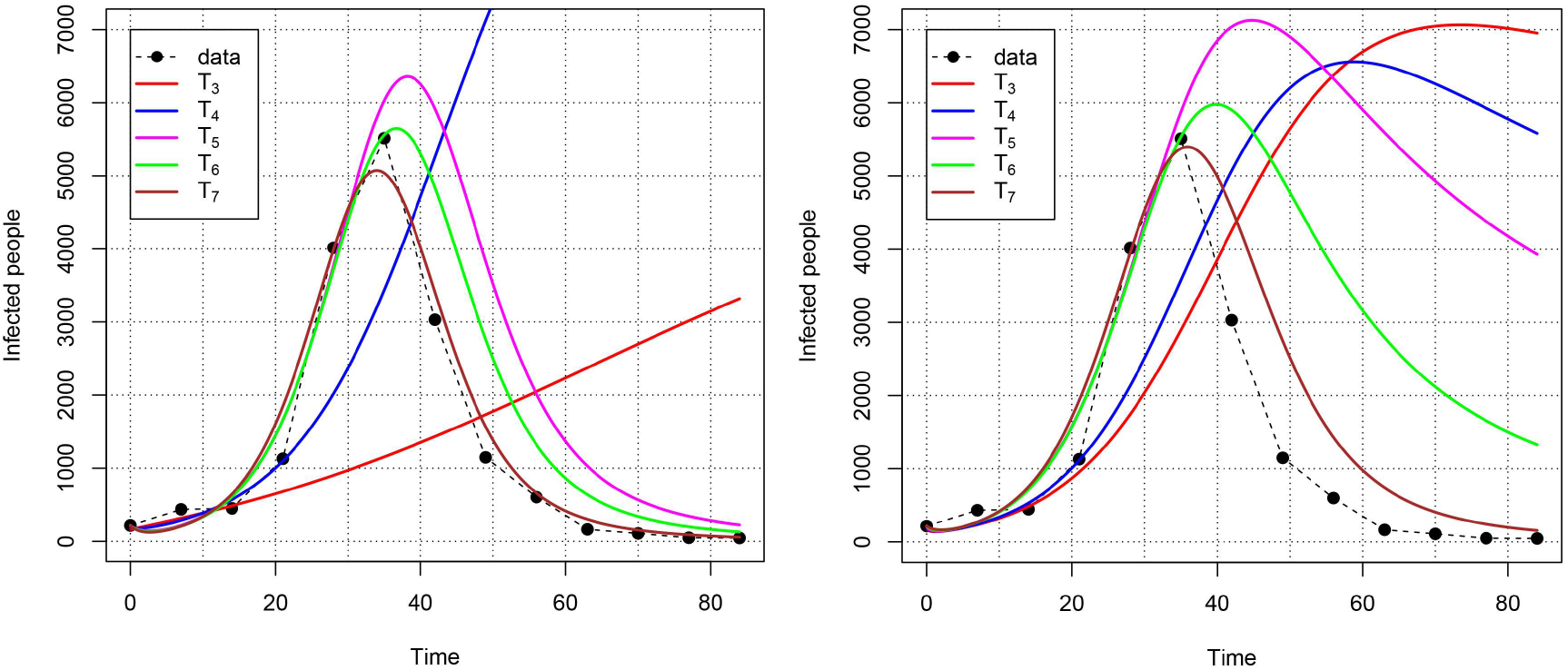
Identification precedure without weights for different number of data for Nishiura (left) and Diethelm (right) fractional models.

**Figure 8.**
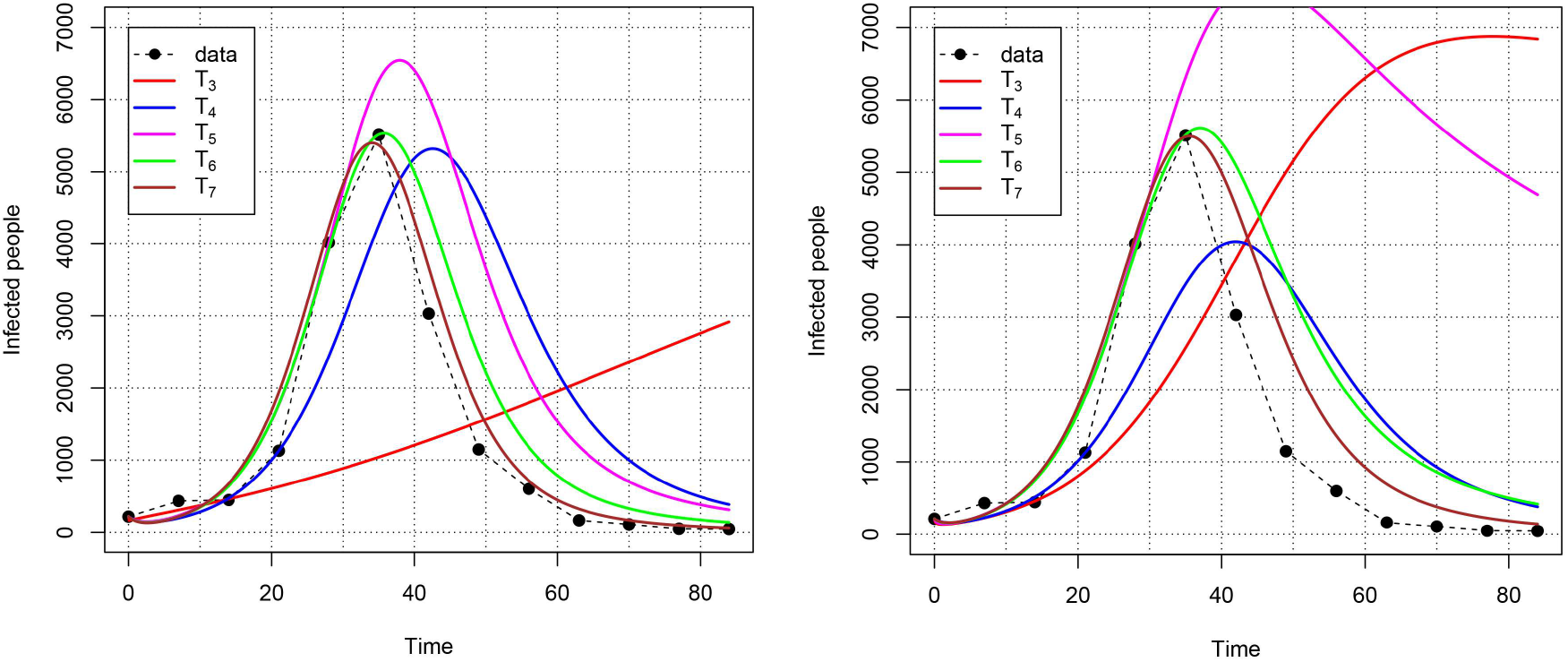
Identification procedure with weights for different number of data for Nishiura (left) and Diethelm (right) fractional models.

As we can see on the above figures, for the initial phase of the epidemic the Nishiura’s model gives faster the more accurate predictions of the epidemic course than Diethelm’s model. The best results can be observed for Nishiura’s model with the weight function, which leads to the conclusion that it is reasonable to enhance the impact on the identification procedure of data from the main course of the epidemic.

## 4 Conclusion and perspectives

In this work, we focus on the global and partial observability of fractional models for complex systems. After defining a formal mathematical framework for the parameter identification problem in FDE models, we introduce a new general algorithm to calibrate partially observable fractional models. This algorithm is particularly suited to life science applications that require preserving key invariant properties between the classic ODE model and its fractional counterpart, as well as between the continuous system and its discrete version used for simulation. In particular, our new method ensures both homogeneous parameter units and solution positivity. Indeed, parameter units are often altered when replacing integer-order derivatives with fractional ones, yet most existing approaches do not address this issue. Solution positivity is also preserved during simulation through a dedicated numerical scheme [27]. Moreover, our algorithm integrates a weighted loss function that accounts for time-dependent data reliability. This method is general and can be applied to a wide range of complex problems in life sciences beyond epidemiology.

To illustrate the capabilities of the method, we consider the modeling of a Dengue outbreak, as such an epidemic concentrates all the challenges mentioned above: (i) system complexity (two types of individuals, four serotypes, seasonal impacts, and memory effects), (ii) partial observability (only newly infected individuals are observed, corresponding to one out of five variables in the model), (iii) data reliability (incidence data are noisy, limited, and often not representative, especially in the early stages of the outbreak), an d (iv) the need to preserve important invariant model characteristics (positivity and parameter units). We thus propose a new fractional homogeneous model for Dengue outbreaks based on the Nishiura model [38], and apply our algorithm to calibrate it using incidence data from a Cape Verde epidemic. A comparative study, applying our algorithm to both the fractional homogeneous Nishiura model and another fractional Dengue model, with and without weighting, shows that our approach improves the fit to data in both cases. Furthermore, the fractional homogeneous Nishiura model appears to provide a simpler and effective alternative to previous approaches for modeling Dengue virus propagation. Finally, our method allows us to determine the minimal amount of data required for parameter identification in order to accurately predict the epidemic peak.

Our study was limited to case of study and use only incidence data. A natural question is now to which extent the fractional exponents obtained for the Cape Verde Dengue outbreak are valuable for any Dengue outbreak in the world. In order to answer this question, we plan to use machine learning techniques [7, 8] adapted to fractional systems on the data base of the Dengue outbreaks in India. Indeed, the Dengue virus is endemic in India and several outbreaks take place each year. In particular, we want to know if the fractional exponent are dependent of the geographical configuration of the place it appears. Another perspective will be to use new types of data to calibrate the model to see if they improve or not the fitting. For instance, we can integrate entomological data regarding mosquito monitoring and control, number of people vaccinated, serological surveys and even human contact patterns from transport flow.

## Data Availability

All data produced in the present work are contained in the manuscript or available on request.

## Acknowledgements

A. Szafrańska thanks the National Science Center for the financial support, under the research project No. 2021/05/X/ST1/00332.

